# Framework for rational donor selection in fecal microbiota transplant clinical trials

**DOI:** 10.1101/19000307

**Authors:** Claire Duvallet, Caroline Zellmer, Pratik Panchal, Shrish Budree, Majdi Osman, Eric Alm

**Affiliations:** Department of Biological Engineering, Massachusetts Institute of Technology, Cambridge, MA 02139; Center for Microbiome Informatics and Therapeutics, Cambridge, MA 02139; Biobot Analytics, Somerville, MA 02143; OpenBiome, Cambridge, MA 02140; Harvard Medical School, MA 02115; The Broad Institute of MIT and Harvard, Cambridge, MA 02139

## Abstract

Early clinical successes are driving enthusiasm for fecal microbiota transplantation (FMT), the transfer of healthy gut bacteria through whole stool, as emerging research is linking the microbiome to many different diseases. However, preliminary trials have yielded mixed results and suggest that heterogeneity in donor stool may play a role in patient response. Thus, clinical trials may fail because an ineffective donor was chosen rather than because FMT is not appropriate for the indication. Here, we describe a conceptual framework to guide rational donor selection to increase the likelihood that FMT clinical trials will succeed. We argue that the mechanism by which the microbiome is hypothesized to be associated with a given indication should inform how healthy donors are selected for FMT trials, categorizing these mechanisms into four disease models and presenting associated donor selection strategies. We next walk through examples based on previously published FMT trials and ongoing investigations to illustrate how donor selection might occur in practice. Finally, we show that typical FMT trials are not powered to discover individual taxa mediating patient responses, suggesting that clinicians should develop targeted hypotheses for retrospective analyses and design their clinical trials accordingly. Moving forward, developing and applying novel clinical trial design methodologies like rational donor selection will be necessary to ensure that FMT successfully translates into clinical impact.

## Introduction

Fecal microbiota transplantation (FMT) is the transfer of gut bacteria through whole stool from a healthy donor to a recipient. FMT has demonstrated high cure rates in recurrent *C. difficile* infection (CDI) across multiple randomized, placebo-controlled trials (Quraishi et al. 2017) and has now entered standard of care for multiply recurrent CDI in European and North American guidelines (McDonald et al. 2018; Cammarota et al. 2017; Surawicz et al. 2013). Beyond CDI, FMT is being explored in range of microbiome-mediated diseases, and has demonstrated promising results in inflammatory bowel diseases (Panchal et al. 2018; Gelfand 2018; Kootte et al. 2017; Osman 2018; Costello et al. 2017; Paramsothy et al. 2017).

Despite these early successes, the underlying mechanism of FMT across all disease indications, including CDI, remains unclear. However, it is generally considered that FMT restores gut microbial community perturbations from a dysbiotic state to a healthy stable state with engraftment of donor strains, or perhaps through other donor-dependent features such as the abundance of non-bacterial components or donor clinical features (Ott et al. 2017; Zuo et al. 2018). However, not all FMT donors are alike: gut microbiota compositions vary within healthy populations in ways that could impact the findings from an FMT trial (Yatsunenko et al. 2012, Wilson et al. 2019). This critical point of microbiome variation within healthy donors is rarely considered in the development of FMT trials (Bafeta et al. 2017; Olesen et al. 2018).

Unlike FMT trials in CDI, where selecting donors based on specific clinical or microbiome profiles does not seem to affect clinical response rates, donor selection is likely to be crucial to trial outcomes in diseases with more complex host-microbiome interplay or distinct disease-associated perturbations (Wilson et al. 2019). Most notably, in a randomized controlled trial (RCT) of FMT for ulcerative colitis (UC) using 5 donors, 78% of patients who achieved remission after FMT received stool from a single donor (Moayyedi et al. 2015). Thus, it is possible that without this single donor, the trial would have returned a negative result. Given the variation in healthy donor microbiomes and donors’ potential impact on clinical efficacy, how should clinicians and investigators select their donors for a clinical trial?

To date, the typical approach for donor selection in FMT trials is to use a single healthy donor or to randomly select multiple donors from a set of screened potential donors (Paramsothy et al. 2017; Kelly et al. 2016; van Nood et al. 2013). However, in clinical indications where successful donors may be rare, such as UC, clinical trials with randomly-selected healthy donors may fail not because FMT is inappropriate for the indication, but because an ineffective donor was chosen. An alternative approach is to expose each patient to multiple donors in order to mitigate the risk of sub-optimal donor selection. In a large RCT of FMT in UC, FMT enemas for a single patient were derived from between three and seven donors with patients receiving multiple donors throughout the 8 week course of treatment (Paramsothy et al. 2017). However, using multiple donors for a single patient may not be feasible or appropriate in many disease indications or clinical trial settings (e.g. single-dose FMT studies). Continuing the practice of randomly selecting donors for FMT clinical trials risks returning false negative trials, stalling the field and delaying the development of novel therapies for microbiome-mediated conditions.

Unlike traditional clinical trials which test well-defined small molecules, FMT trials test the donor microbiome, which is variable (Olesen et al 2018). Fortunately, the emergence of large, stool banks with multiple pre-screened healthy donors captures some of this variability and makes it available for use in FMT trials. These stool banks thus open the possibility of selecting donors rationally during the FMT clinical trial design phase, enabling clinicians to choose from among a large pool of eligible donors for donor samples which have specific desirable characteristics. Coupled with expanded access to genome sequencing technologies and publicly available microbiome sequencing datasets, rational donor selection is feasible and presents a unique opportunity to advance the research methods of this nascent field.

In this paper, we present a framework to guide donor selection for FMT trials. The mechanism by which the microbiome is hypothesized to be associated with a given indication should inform how donors are selected for FMT trials, and we describe different disease models which may underlie microbiome-mediated conditions (Figure 1). We describe strategies to rationally select donors for each type of disease model, and provide examples based on previously published FMT trials and ongoing investigations. Finally, we discuss limitations of performing discovery-based retrospective research after an FMT clinical trial concludes. To our knowledge, this is the first description of a comprehensive framework for rational donor selection in FMT trials.

**Fig 1:**
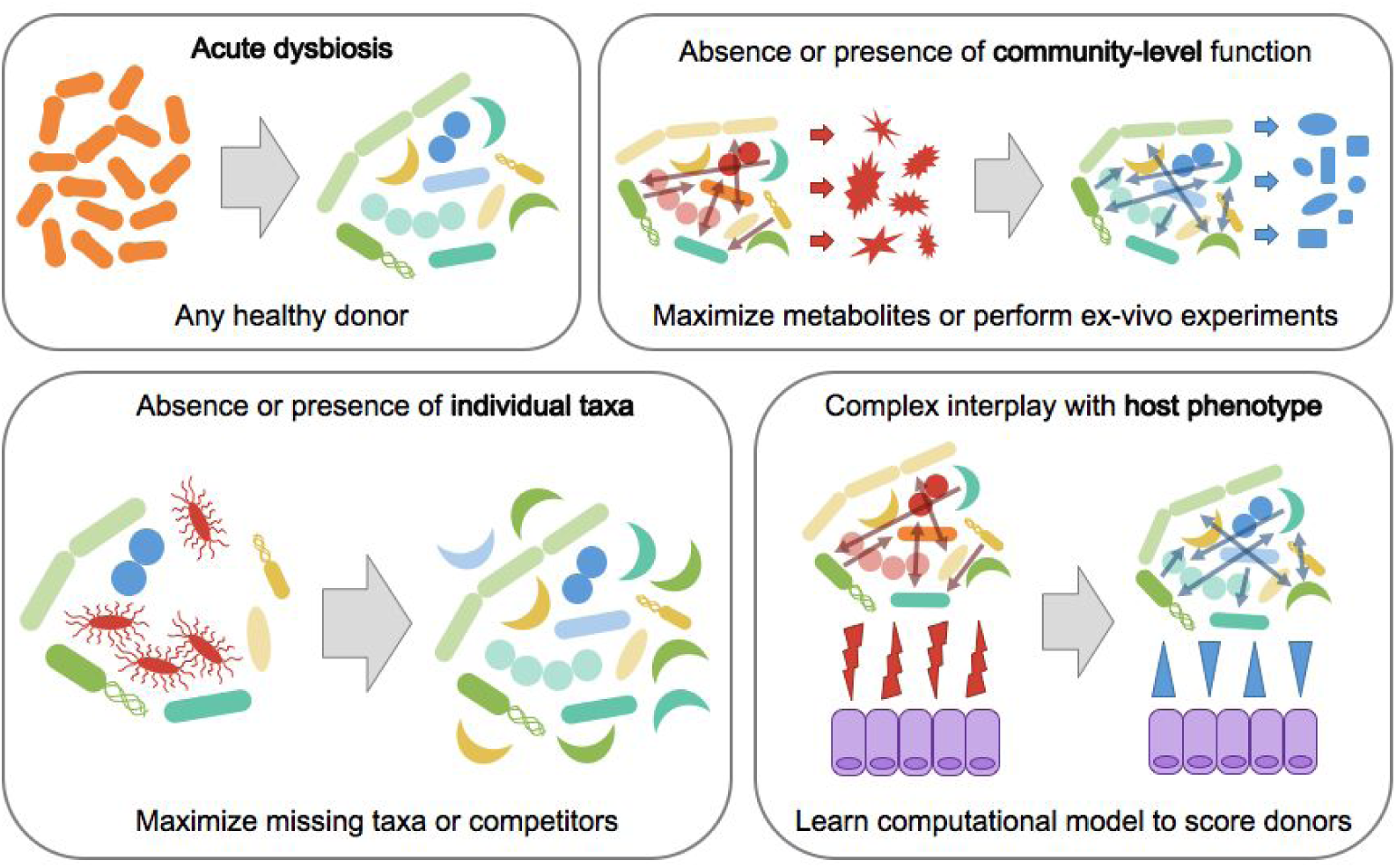
overview of the different models of microbiome-mediated disease and associated donor selection strategy. In cases where the underlying model is unknown, a variety of donor selection approaches could be employed to potentially identify which disease process(es) may be involved.

### Framework for rational donor selection

FMT trials are pursued when research or clinical experiences suggest that the microbiome may be causing or exacerbating a disease. Here, we propose four different models which may underlie microbiome-mediated etiologies and their corresponding rational donor selection strategies (Figure 1). Ultimately, it is up to each individual clinician-researcher to use published cross-sectional studies, mechanistic investigations in model organisms, and their own clinical experience treating patients to determine which of these model(s) are relevant in their specific case. Additionally, logistical considerations will be important factors in making the final donor selection regardless of which strategy is pursued. For example, clinicians should ensure that the pool of donors that they are screening have enough material to sustain the required number of FMTs for their entire trial. Finally, this framework represents an approach for optimizing the success of FMT clinical trials given that a clinician is already pursuing a trial, and is not intended to be used for deciding whether or not an FMT trial should be pursued in the first place.

Most of the donor selection strategies described below can be modified to incorporate matching between patients and donors. More specifically, donors can be tailored to individual patients to specifically make up for the unique taxonomic or functional deficiencies in that patient’s microbiome (Wilson et al 2019). With the increasing amount of microbiome data available from published FMT trials, we encourage collaborations between clinicians and bioinformaticians to analyze these data in order to generate or perhaps even confirm the validity of potential donor selection strategies before selecting one (Figure 2). Finally, the strategies presented here should also be combined with adaptive clinical trial designs to further increase the probability of having a successful FMT trial (Olesen, Gurry, and Alm 2017).

**Fig 2:**
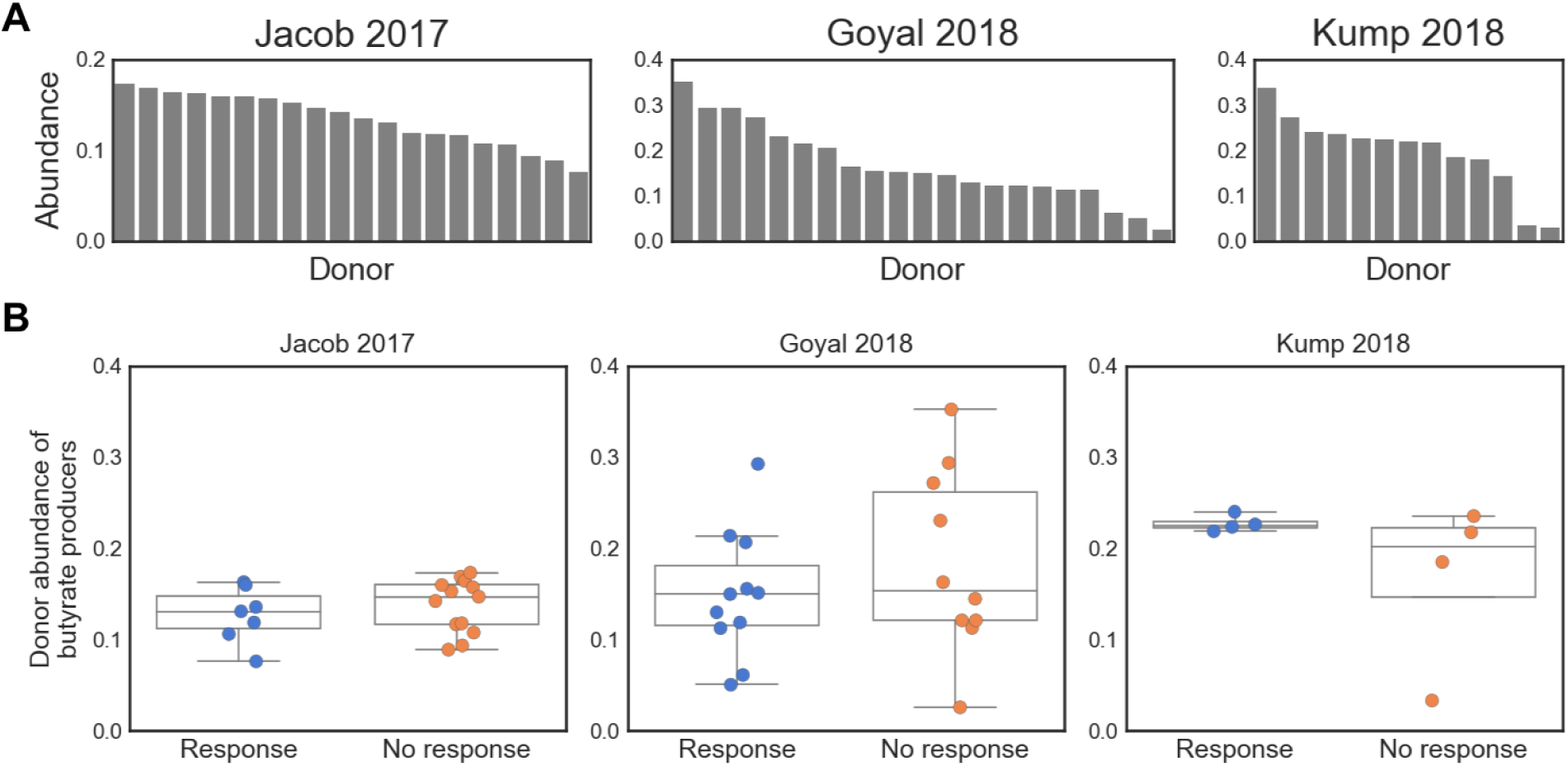
Case study in IBD: select donors based on abundance of butyrate producers? (A): abundance of butyrate producers in each study’s donor samples, showing that healthy donors exhibit a range of abundances. Butyrate producers were identified at the genus-level (see Methods). (B): abundance of butyrate producers in donor samples, stratified by respective patient’s response. Patient response was defined as in the original publications: Jacob 2017, response at 4 weeks (ΔMayo score ≥ 3 and bleeding subscore ≤ 1); Goyal 2018, response at 6 months (decrease of 15 points in PUCAI); Kump 2018, response at day 90 (reduction of the total Mayo score by ≥ 3 points). Patient response is not significantly associated with the abundance of butyrate producers in their respective donor sample (p > 0.2 for all response vs. no response comparisons, t-test).

In all cases, ensuring patient safety through carefully screening donors and following all regulations relating to pursuing an FMT trial is of paramount importance and supersedes any other considerations regarding donor selection. While FMT has shown promise and is defined in some clinical guidelines as standard of care for multiply recurrent CDI (McDonald et al. 2018; Cammarota et al. 2017; Surawicz et al. 2013), it is not yet officially approved by the US FDA. Thus, clinicians should ensure that their trial has secured all ethical and regulatory approvals before proceeding (FDA 2013). Additionally, only healthy donor stools should be considered in the rational selection process. Donors should be evaluated through assessments that exclude not only known pathogenic risk but also co-morbidities and risk factors for indications that are associated with the microbiome but where causality is unknown. Following two serious adverse events in immunocompromised individuals receiving FMT, the FDA recently implemented additional restrictions regarding screening requirements for donor stools (FDA 2019). As our understanding of the risk factors related to FMT increases, donor screening criteria should be updated to reduce the likelihood of serious adverse events in patients, regardless of where donor stool is sourced and whether rational donor selection is performed. For example, OpenBiome, the first public stool bank, has implemented a rigorous screening protocol to exclude donors with any of a variety of potential clinical and infectious risk factors (OpenBiome 2019).

### Models of microbiome-mediated disease

#### Acute dysbiosis

An acutely dysbiotic gut microbial community is broadly dysfunctional and can no longer maintain the health of the host. For example, in the case of recurrent *Clostridium difficile* infection (rCDI), a disturbed microbial community is unable to prevent colonization by or overgrowth of the pathogen, leading to recurrent overgrowth of *C. diff* and clinical symptoms (Britton and Young 2014). Acute dysbiosis has also been described with the “Anna Karenina principle”: all healthy microbiomes are alike but dysbiotic communities are all dysbiotic in their own ways (Zaneveld et al 2017). In this view of acute dysbiosis, microbial communities respond stochastically to stressors, resulting in dysbiotic communities which are characterized by increased variability rather than deterministic shifts to precise community type(s) (Zaneveld et al 2017).

In this model, the host simply needs to return to a “healthy” microbiome and thus choosing any healthy FMT donor should be sufficient to induce clinical improvements. Because there is no specific disease-associated microbial community and deviation from health is instead the more important factor, simply replenishing the microbiome with a healthy configuration should be sufficient. Indeed, FMT trials have demonstrated that rCDI can be effectively treated by almost any choice of healthy donor (Osman et al. 2016). In this case, researchers should consider how they define a “healthy” microbiome and how they will ensure clinical efficacy, for example through engraftment of the transplanted healthy communities.

### Absence or presence of individual taxa

#### Absence of beneficial taxa

In other cases, perhaps a disease is being caused or exacerbated by the lack of certain specific microbes, and replenishing these few taxa would be sufficient to restore the host to health (Wilson et al 2019). For example, Hsiao et al. showed that a single microbe, *R. obeum*, restricted infection by *V. cholerae* through quorum-sensing-mediated mechanisms (Hsiao et al. 2014). Surprisingly, non-communicable diseases may also fall into this model: a single strain of *Lactobacillus* was sufficient to ameliorate salt-induced hypertension in mice, and follow-up studies indicate that similar mechanisms may be involved in salt-sensitive high blood pressure in humans as well (Wilck et al. 2017).

In these cases, the donor selection strategy should focus on maximizing the probability of engraftment of the beneficial taxa. In cases where the unique taxa are not specifically known or are rare members of the human microbiota, many healthy donors could be pooled together or a donor with a high alpha diversity could be selected in order to maximize the probability that the transplanted sample contains the necessary taxa (Wilson et al 2019). However, pooling donors may increase the risk of adverse events and should be pursued with caution. If the missing microbes are known and well-characterized, on the other hand, researchers can screen their pool of potential donors to find the sample with the highest abundance of these taxa.

#### Presence of harmful taxa

Rather than being characterized by the absence of individual bacteria, perhaps a disease is instead mediated by the presence or overabundance of specific microbes, and removing these bacteria in a targeted fashion could lead to improvements in disease progression. For example, *Fusobacterium* has been found to be more abundant in colorectal cancer patients, specifically enriched in the tumors themselves (Kostic et al. 2013). Multiple groups have identified mechanistic associations between *Fusobacterium*, inflammatory transcriptional signatures, and tumor growth in mouse and human models of colorectal cancer, pointing to a causal role for *Fusobacterium* in colorectal cancer progression (Kostic et al. 2013; Rubinstein et al. 2012). Recent work has found that treating tumors with antibiotics slows tumor progression, further confirming these causal associations and pointing toward potential microbiome-based therapeutic interventions (Bullman et al. 2017).

Removing and replacing these bacteria should be the goal of FMT in cases where this disease model applies. This can be achieved by first removing the harmful bacteria (e.g. via antibiotic treatment) with follow-up FMT to re-establish a healthy community that prevents their re-colonization. In all cases, donors should be screened to exclude any samples which contain the harmful bacteria. Donor samples can then be selected based on the abundance of bacteria which are known to out-compete the harmful taxa. Competitors can be identified by searching the microbiology literature to identify bacteria which live in the same niche or which have been experimentally shown to directly out-compete the undesirable taxa, or they can perform these competition assays themselves. If resources to perform competition assays are not available and the literature is sparse, researchers can also mine existing microbiome data to find bacteria which consistently anti-correlate with the harmful taxa, and choose donor samples with a high abundance of these putative competitors (Friedman and Alm 2012).

#### Patient matching

Taxa-based donor selection strategies are particularly amenable to patient-matching, when both patient and donor microbiome data are available prior to the start of a trial (Wilson et al 2019). For example, if one patient is completely missing some of the beneficial taxa but not others, then these taxa can be weighted more heavily in the donor selection process. The phylogenetic relationships between donor and recipient taxa could also be incorporated into donor selection: if a patient already has many bacteria which are closely phylogenetically related to known competitors of some of the harmful bacteria, then competitors of the other harmful bacteria can be upweighted in the donor selection process. Similarly, if patients already have taxa which are already filling certain niches important for health, the taxa which fill those same niches can be downweighted in donor selection.

#### Case study: Inflammatory Bowel Disease

An example where the “missing taxa” model may be applicable is in inflammatory bowel disease (IBD). Butyrate has long been associated with IBD (Wilson et al 2019, Scheppach et al. 1992), and recent case-control and longitudinal studies point to a consistent lack of butyrate-producing bacteria in patients with IBD (Duvallet et al. 2017; Schirmer et al. 2018). Furthermore, preliminary FMT trials in IBD have been marked by variable efficacy, both between trials and between donors within individual trials, suggesting that donor microbiome characteristics may be associated with FMT response (Kump et al. 2018; Moayyedi et al. 2015). These results indicate that IBD may benefit from rational donor selection approach, and that donors with high abundances of butyrate-producing organisms may yield higher FMT response rates than randomly selected donors.

Given the availability of microbiome data from completed FMT studies, a clinician leading an IBD FMT trial may elect to perform rational donor selection based on the “absence of beneficial taxa” disease model. To illustrate this process, we re-analyzed microbiome data from three completed IBD FMT trials which provided publicly available sequencing data for patient and donor samples (Kump et al. 2018; Goyal et al. 2018; Jacob et al. 2017). We selected butyrate-producers based on their genus-level taxonomy, using a simple heuristic from Vital et al. 2017 (see Methods). Donors in the three studies exhibited a range of total abundances of butyrate-producing bacteria (Figure 2A, Supplementary Figures 1-3). However, the abundance of butyrate producers in the donor stool was not associated with recipient patients’ clinical responses (Figure 2B) and we also found no association with response when matching donor abundances with their respective patient’s original abundance of butyrate producers (Supplementary Figure 4). This illustrative case study shows that selecting donors based on the abundance of butyrate producers may not yield improved clinical trial outcomes in IBD, and more broadly highlights some of the challenges involved in performing rational donor selection in a real-world context.

Despite their accessibility, taxonomy-based approaches are limited in their ability to identify functional or strain-level associations. For example, in this analysis, we were unable to identify any of the Eubacteria taxa which are major contributors to SCFA production in the human gut (Vital et al. 2017, Louis et al, 2010). More complex methods to identify butyrate producers (e.g. using more finely resolved taxonomy, phylogenetic-aware methods and/or metagenomics data) could be used in the next iteration to develop a donor selection strategy, if these data are available to clinicians. Another approach, discussed below, is to select donors based on functional community assays and direct measurement of butyrate production rather than microbial taxonomies alone.

### Community-level functionality

Some microbiome-associated diseases may not be addressable by replenishing the patient with a generically healthy community or by targeting individual taxa, and may instead be mediated by the microbiome through a community-level function. Here, there may not be a consistent disease-associated microbiome across patients in terms of taxonomic composition, but patients may be characterized by having microbiomes which are similarly missing or enriched in some core functionality. This model may also apply to conditions where there are consistent disease- or health-associated taxa, but in which their collective functioning is the more important mediator of disease. The IBD case study described above may reflect this situation: although depletion of butyrate producers is strongly associated with IBD throughout the literature, a successful donor selection strategy may need to consider butyrate production directly rather than through the proxy of taxonomy (Wilson et al 2019; Duvallet et al. 2017; Schirmer et al. 2018).

#### Missing community-level function

In the case where a community-level function is missing from patients’ microbiomes, the goal of FMT should be to replace the deficient community with a beneficially functional microbiome. Here, it is important that a single donor with an intact microbial community is used, rather than a mixture of donors which may not yield the desirable community composition at steady-state after FMT. To choose a donor, molecules which can serve as proxies for the metabolic output of the microbial community can be measured directly in donor stool, and donors can be selected based on the abundance of these molecules.

Like IBD, hepatic encephalopathy (HE) is an example where community functionality is likely more relevant to FMT outcome than specific taxa. A previous trial in HE (Bajaj et al. 2017) rationally selected their single donor by maximizing the abundance of *Lachnospiraceae* and *Ruminococcaceae*, taxa which were were previously found to be depleted in HE patients based on cross-sectional microbiome data. The clinical trial was a success, but it remains unclear from this trial whether the donor’s strains engrafted in the patients post-FMT and whether this played any role in the successful FMT responses. The exact mechanisms of action of these strains remain unknown, though both bacterial families are known short chain fatty acid producers (in particular butyrate) (Vital et al. 2017). Recent studies have more directly implicated deficiencies in the production of short-chain fatty acids (SCFAs) and secondary bile acids as being important in liver cirrhosis and subsequent complications such as HE, suggesting that community-level functioning may be a more important driver of FMT response. Thus, HE may be a case in which function-based donor selection can be employed. To illustrate this process, we analyzed stool metabolomics data from 83 OpenBiome donors and used this data to rank them based on their estimated production of SCFAs and secondary bile acids (Figure 3; Poyet, Groussin, Gibbons, et al. 2019). As in the IBD case study (Figure 2A), we found that donors exhibited a range of values for our metabolites of interest (Figure 3A and C). We ranked donors based on their amounts of the three measured SCFAs (butyrate, isovalerate, and propionate) and on their bile acid conversion rates, approximated as the ratio between the total amounts of primary and secondary bile acids (Figure 3B and D, see Methods). With this process, we were able to identify four donors who were in the top 25% of all donors for both metrics (Figure 3E). In a real FMT trial, a clinician would then work with their stool bank to ensure that these donors were still active and/or had enough material to fulfill the full trial, or alternatively request that donors with a similar range of SCFAs and secondary bile acid conversion be provided.

**Fig 3:**
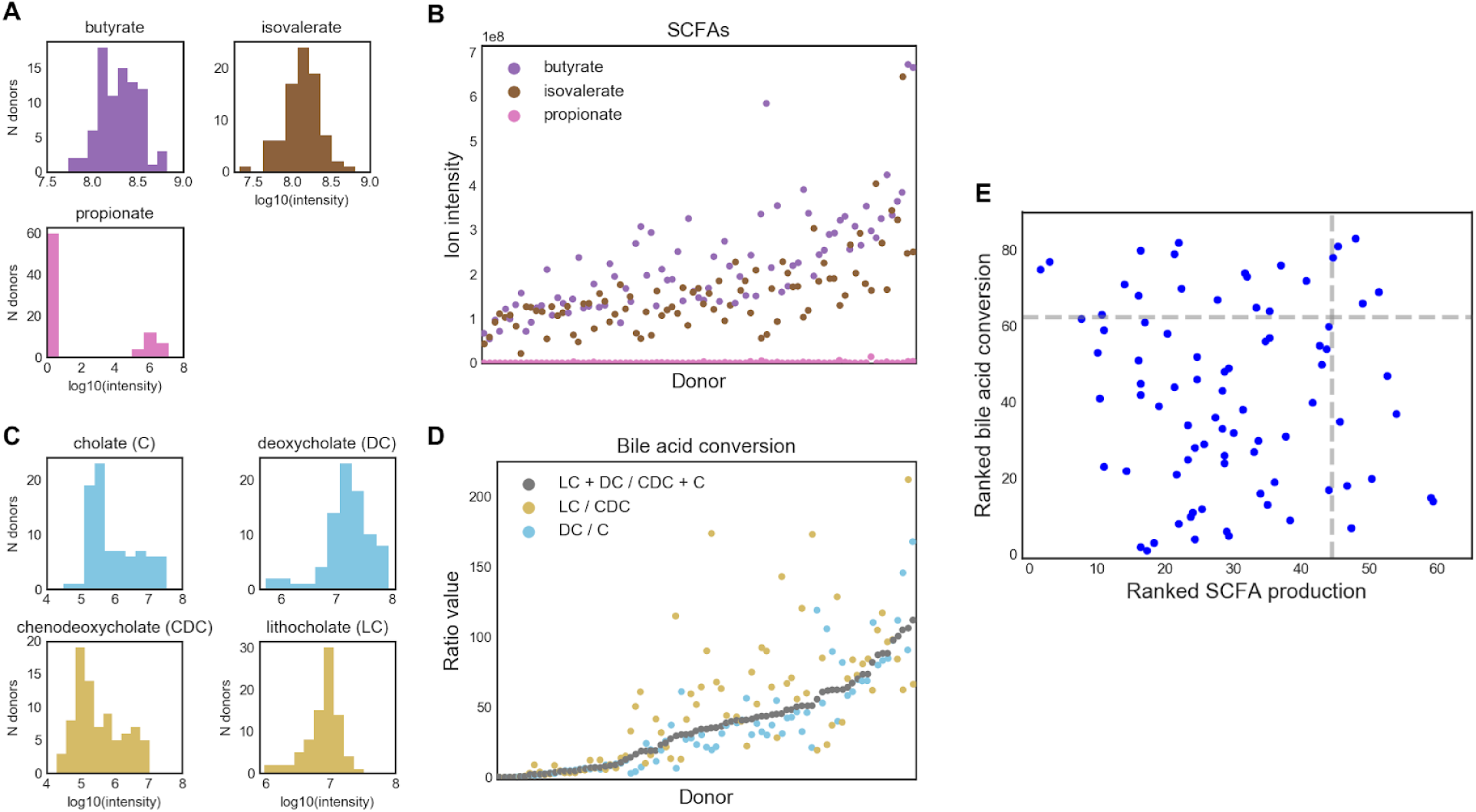
Case study in liver cirrhosis: selecting donors based on community function by mining stool metabolomics data. (A) Distribution of SCFAs in all donor stools. (B) Abundance of each SCFA per donor, ranked by average SCFA abundance. (C) Distribution of bile acids in all donors. Primary bile acids are in the left column, secondary bile acids are in the right column. Bile acids are colored according to pathways. (D) Bile acid conversion ratios in each donor, ranked by the ratio of total secondary to primary bile acids. (C: cholate; DC: deoxycholate; CDC: chenodeoxycholate; LC: lithocholate) (E) Donors ranked on both SCFA production and bile acid conversion allows for the selection of five donors which are in the top 25% for both of these metrics. These donors could be followed up with for use in a rationally-designed liver cirrhosis FMT trial.

While measuring metabolites in stool as a proxy for community production will likely be an improvement over taxonomy-based approaches in most cases, these measurements are also complicated by potential host effects. For example, different hosts may absorb these molecules at different rates, and so measuring them in stool may not be an accurate reflection of each donor community’s productive potential. Additionally, community function may depend on non-biologically relevant factors like the donor’s diet and time that they provided their sample. As an example, bile acid production spikes after meals (Hofmann 1989), so the amount of bile acids measured in a given stool sample may reflect the amount of time since the donor last ate rather than their actual microbial community’s functional production of these molecules. If clinicians have access to sufficient resources, a better way to screen donors may be to perform *ex vivo* assays, in which each donor sample is homogenized and provided with the substrates (e.g. fiber) needed to produce the desirable output (e.g. short-chain fatty acids like butyrate). In this way, the donor community function can be measured directly (Wang and Gibson 1993; Chen et al. 2017).

#### Overactive function

A disease may also be mediated by an overactive microbiome doing something harmful to the host. For example, TMAO produced by the microbiota contributes to atherosclerosis (Koeth et al. 2013; Wang et al. 2015). Here, the goal of FMT should also be to replace the patient’s microbiome with a beneficially functional community, but the donor selection strategy may attempt to identify communities in which the harmful function is completely absent or which produces an inhibitor of the harmful microbe-derived molecule (Wang et al. 2015).

### Microbiome-associated host phenotypes

Diseases with more complex etiologies may not have a direct taxonomic or functional association with the microbiome but instead be related through some intermediate host phenotype which needs to be improved or corrected. For example, severe acute malnutrition has been associated with a gut microbiota which is not fully mature, with mouse experiments suggesting that this association may be causal (Blanton et al. 2016; Subramanian et al. 2014). Other studies have shown a relationship between gut microbiome, immune development, and development of autoimmune conditions later in life (Stokholm et al. 2018; Cox et al. 2014; Kostic et al. 2015). These relationships may have mechanistic explanations which are not directly measurable from donor or patient stool (e.g. immunogenicity of bacteria, ability of bacteria to digest the host’s mucus) but which can nevertheless be inferred from existing data and used to select potential donors.

For these more complex cases, models can be trained from existing datasets to learn the community signatures linked to the disease-associated phenotype. In some cases, it may be possible to develop computational models which directly predict the phenotype of interest. For example, Stein et al. developed a model to predict the induction of regulatory T-cells by microbial communities (Stein et al 2018). In other cases with few known mechanistic models, machine learning algorithms can be trained on multiple cross-sectional datasets to identify complex signatures that reproducibly distinguish patients from healthy controls. These models can then be applied to score potential donors, and the donor with the “most healthy” score may be chosen for a trial.

### Little understanding of underlying disease model

In some conditions, there may not be enough understanding of the underlying microbiome-based etiology to inform donor selection in an FMT trial. It may also be the case that there are no existing datasets on which to train models, existing datasets are not sufficiently powered to distinguish the different potential underlying models, or logistical considerations constrain a clinician’s ability to select specific donors for their trial. In these cases, we recommend selecting different healthy donors, employing an adaptive clinical trial design in which donors are cycled after they have clinical failures (as described previously in Olesen, Gurry, and Alm 2017), and performing retrospective analyses to answer targeted hypotheses which were developed during the clinical trial design process.

#### Cycling healthy donors in adaptive trials

As donors change through the course of an adaptive trial, clinicians may elect to select their donors randomly or to more rationally cycle through donors (Olesen, Gurry, Alm 2017). “Differently healthy” donors may be selected through a combination of donor selection strategies to represent the different underlying disease-associated models described above. Clinicians may be able to test whether a biological mechanism is involved by strategically selecting donors and testing whether one type of donor led to better outcomes than the others. For example, if the hypothesized biological mechanism relates to butyrate production, healthy donors with high and low levels of butyrate production could be used in the FMT arm, and retrospective analyses may be able to determine whether there is an association between donor butyrate levels and FMT outcome. However, since clinical trials prioritize clinical efficacy over secondary outcomes like mechanistic insights, few exploratory FMT trials will likely have the sample size and power necessary to conclusively perform such analyses.

Donors may also be selected to span the range of “healthy” microbiomes in a given population. For example, clinicians may pick a “median” healthy donor who is similar to existing healthy reference microbiomes (HMP Consortium 2012, Halfvarson et al 2017), or simply based on the presence or abundance of certain consistently “core” health-associated bacteria (Shade and Handelsman 2012; Duvallet et al. 2017). In a similar vein, “healthy” donors can also be chosen based on their distance from disease-associated microbiomes, as opposed or in addition to similarity to health. Published case-control datasets can be used to identify donors with communities which are farthest away from the median or average diseased patient. These datasets can also be mined to identify taxa which are consistently disease-associated, and which should be minimized or perhaps even absent in the potential donor. Pairing rational donor selection with adaptive trial designs may eventually yield insight into the underlying model mediating the disease of interest if certain types of “healthy” donors consistently perform better at treating patients than others.

#### Discovery-based retrospective analyses

In these exploratory FMT clinical trials, discovering microbiome characteristics which are differentially associated with FMT response may be a valuable secondary endpoint (Olesen et al. 2018), identifying characteristics of good donors and informing donor selection strategy for future trials (Kump et al. 2018, Jacob et al. 2017, Fuentes et al. 2017). Furthermore, companies attempting to develop microbiome-based therapeutics may use FMT trials to discover the key bacteria which mediate FMT response in order to include these in their microbial cocktail product. However, exploratory FMT trials tend to enroll few patients, limiting the potential power of retrospective analyses of individual trials to find associations between the microbiome and FMT response.

We performed a simulation to determine the likelihood of a retrospective analysis to identify donor-derived bacteria associated with different patient responses to FMT. We performed this simulation for multiple FMT trial set-ups and outcomes (i.e. number of FMT responders and non-responders). We used existing microbiome datasets to model different effect sizes, where we use “effect size” to mean the number of bacteria which are differentially abundant in donor samples given to patients who did and did not respond to FMT. Case-control datasets were used to model the microbiome data and various effect sizes, with a large effect represented by an infectious diarrhea dataset (Schubert et al. 2014), a medium effect represented by colorectal cancer (Baxter et al. 2016), and a weak effect represented by obesity (Goodrich et al. 2014). For each of these datasets, we identified the top ten most differentially abundant bacteria in the overall population as representative of “key mediating bacteria” (see Methods). Next, we simulated different trials, varying the numbers of patients in the FMT arm and the FMT response rates (i.e. proportion of patients which were FMT responders, represented by sampling from the “case” patients, vs. non-responders, represented by sampling from the “control” patients). We subsampled patients according to these parameter settings, identified differentially abundant genera, and compared these to the top ten genera identified from the entire datasets (Figure 4).

**Fig 4:**
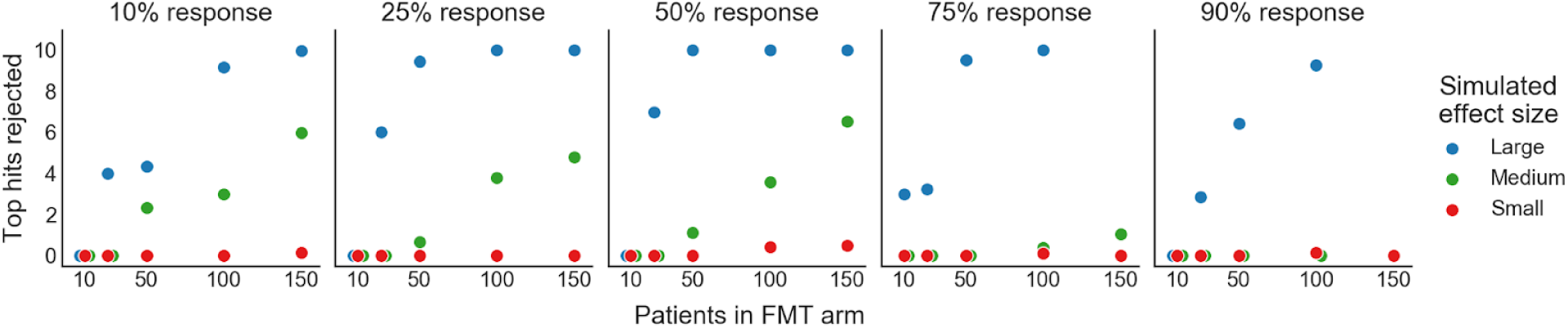
Power simulation: most FMT trials are not powered to discover individual taxa mediating FMT response. Plots showing how many of the 10 most “truly” differentially abundant genera would be recovered as significant under different FMT study designs. Each panel represents a different FMT response rate (i.e. percent of patients in the responder vs. non-responder group). The effect size (i.e. number of genera which are differentially abundant in FMT responders vs. non-responders) was simulated by using three different case-control microbiome datasets. A large effect size is modeled by the effect of diarrhea on the microbiome, medium by colorectal cancer, and small by obesity. The top 10 “true” differentially abundant genera were identified by calculating their signal-to-noise ratios in the full original dataset (i.e. mean difference divided by the standard deviation). (See Methods).

In cases where the microbial signature for FMT response is expected to be large (i.e. the difference in donor stools given to FMT responders vs. non-responders is as large as the effect of diarrhea effect on the microbiome), we found that small FMT trials would recover most of the top hits in the majority of cases. The power to detect associations decreased as FMT response rates became less balanced (i.e. response rates different from 50%), and in these cases trials would need to include up to 50 patients in the FMT arm to recover the key mediating taxa. For both medium and small effect sizes, however, prohibitively large FMT arms would be needed to recover most key mediating taxa regardless of FMT response rate. We found that when the microbial signature for FMT is equivalent to the effect of diseases like colorectal cancer on the microbiome, at least 100 patients are needed in the FMT arm to recover at least half of the most truly differentially abundant genera for most FMT trials. This simulation was performed at the genus-level, and underestimates the power needed to identify associations at finer taxonomic levels (e.g. OTU, ASV, or strain-level). In cases where strain-level biology is involved, FMT trials will need even more patients to be powered to find significant associations in retrospective analyses.

These results suggest that successful secondary analyses of microbiome data from FMT trials will require either very large FMT arms, investigating more targeted hypotheses, meta-analyses, or additional sample collections. For example, clinicians may consider pairing donor and patient samples or collecting longitudinal patient samples to increase power to make discoveries (Fuentes et al. 2017, Chu et al. 2019). They may also consider testing specific hypotheses developed before the trial, such as comparing the total abundance of butyrate producers between FMT responders and non-responders, or performing functional assays to measure specific metabolites thought to be associated with FMT response (Fuentes et al. 2017). On the other hand, researchers wishing to identify the key taxa to include in an FMT drug may consider pursuing clinical trials in which identifying these taxa is the primary endpoint, and power them accordingly. In all cases, making microbiome sequencing data and associated patient-donor matching and clinical response metadata publicly available will allow for future meta-analyses that will have more power to make discoveries (Duvallet et al 2017).

## Discussion

The framework presented here encourages clinicians to leverage their clinical experience, existing microbiome research and published datasets, the increasing availability of screened healthy donor stools, and partnerships with bioinformaticians to more efficiently translate microbiome-based interventions into clinical impact. Clinicians can apply their existing knowledge and *a priori* hypotheses to determine which microbiome-mediated disease model may underlie their indication of interest, and then select donors accordingly. By rationally choosing donors during the FMT trial design, clinicians will increase the likelihood of successful FMT trials in diseases in which donor heterogeneity affects patient response. Our power simulation analysis also suggests that specific plans for retrospective analyses of the microbiome data generated should be developed during trial design, with targeted hypotheses of interest and sample collection plans tailored accordingly. Otherwise, exploratory analyses of single FMT studies are unlikely to make new discoveries. Paired with adaptive clinical trial designs, FMT trials with rationally-selected donors will become an important tool in advancing translational microbiome research and clinical treatment to improve and save patient lives.

As a conceptual framework for approaching rational donor selection in practice, this work has significant limitations. Many logistical and scientific considerations (e.g. FMT delivery mode, organism survival during FMT prep and delivery, etc) involved in performing FMT trials are not addressed in this framework, and should be considered during the trial design in addition to donor selection. FMT trials are also inherently limited by the material that they use: donor stool. It is unclear to what extent stool will be sufficient to address diseases in which regional differences in gut microbiota composition are involved (e.g. mucus-associated microbes implicated in IBD), and to what extent analyzing fecal samples provides a representative view of the entire GI physiology. Furthermore, while amplicon sequencing may be a good first-pass analysis when more extensive functional data (metagenomics, metabolomics, functional assays) is not available, it is limited in its ability to provide insight on functional or strain-level associations. Further work will be required to address which measurement modality is optimal for performing rational donor selection in exploratory FMT clinical trials, while recognizing that these pilot studies are often constrained for resources.

As FMT-specific clinical trial design methodologies become more developed, many additional questions will need to be addressed. Some of these key questions relate to choosing healthy donors: what defines a “healthy” donor, and when and how should that definition change? Some screening criteria, like excluding certain known pathogens, will apply to all donors. Beyond these, however, there is little consensus on what defines a “healthy” microbiome. Consider a clinician carrying out an FMT trial in an African setting: given that healthy Africans from across the continent are known to have more *Prevotella* than North Americans, it might be advisable to source donors locally to better match the expected healthy state of the patients (Ou et al. 2013; Yatsunenko et al. 2012; De Filippo et al. 2010). But what if the local population has higher asymptomatic colonization rates of undesirable bacteria like opportunistic pathogens, resulting in usual screening criteria excluding many or all potential donors: should the criteria be adapted to allow for local donors, or should donors be sourced from a foreign stool bank whose donors may not match the local population? To answer this question, more research is needed to understand differences between healthy microbiomes globally and their clinical implications (Bello et al 2018; Rabesandratana 2018). Regardless, ensuring patient safety in light of our evolving understanding of the microbiome and FMT is critically important to the success of the field. Identifying criteria that should be applied to all donor stools to ensure the safety of FMT will require balancing stringent screening requirements while also allowing for a large enough pool of healthy donors to facilitate successful trials.

On the patient side, comorbidities, lifestyle, and dynamic disease manifestations present additional challenges and opportunities to improve donor selection and FMT clinical trial designs. How should comorbidities be incorporated into donor selection? Patients with multiple disease processes may be dominated by one disease model or may exhibit a combination of models, perhaps affecting which donors would be optimal for their specific case. For example, a person whose condition involves both acute dysbiosis and community-level dysfunction might respond well to any healthy donor, or may require a more complex combination of total community replacement along with enrichment for community function. Relatedly, diseases that change manifestations over time may benefit from employing different donor selection strategies over the course of a longitudinal FMT trial. Additionally, although there have been no serious adverse events related to FMT material in either clinical practice for rCDI or in clinical trials across adults or pediatrics, could some donors further reduce the probability of adverse events in at-risk patients? Finally, how should other sources of heterogeneity like lifestyle, diet, and medication usage be incorporated into rational donor selection? In cases where FMT is combined with other microbiome-targeted interventions like prebiotics or dietary changes, could some donors have synergistic effects with these paired interventions and lead to greater clinical success?

To ensure that FMT reaches its full potential to improve and save patient lives, clinicians should think critically about how their FMT trials can be designed for maximal impact. Applying new approaches like rational donor selection will decrease the number of false negative clinical trials which fail even though they could have succeeded with a more optimal donor. Furthermore, by developing targeted hypotheses, post-trial analysis plans, and associated sample collection schema alongside the core FMT trial design itself, the number of basic scientific discoveries that are made from each trial will significantly increase. As FMT expands beyond rCDI and microbiome-based therapeutics are developed to target a range of diseases, novel methods and approaches tailored to the unique challenges and opportunities presented by FMT will be critical to ensuring the advancement of translational microbiome science and beneficial impact on patient lives.

## Methods

### Microbiome data processing (IBD case studies)

Raw fastq data files were downloaded from the European Nucleotide Archive using the following accession numbers: Jacob et al 2017, PRJNA388210; Goyal et al. 2018, PRJNA380944; and Kump et al. 2018, PRJEB11841. All data was processed using QIIME 2 (v. 2018.6.0, Bolyen et al. 2018). Briefly, data was imported into QIIME 2 as paired-end (Kump et al. 2018; Jacob et al. 2017) or single-end (Goyal et al. 2018) data, filtered based on sequence quality with ‘quality-filter q-score’, and denoised with deblur using ‘deblur denoise-16S’ (Amir et al. 2017). Representative sequences were assigned taxonomy using ‘feature-classifier classify-sklearn’ with the GreenGenes-trained Naive Bayes classifier provided by QIIME 2 (gg-13-8-99-nb-classifier.qza) (Bokulich et al. 2018.). All data was exported to tab-delimited format and analyzed in Python 2.7.6. We converted raw read counts to relative abundances by dividing each value by the total reads per sample, and collapsed ASVs to genus level by summing their respective relative abundances.

### Quantifying abundance of butyrate producers

We identified butyrate producers at the genus-level based on the analysis performed in Vital et al. 2017. These taxa were detected in >70% of individuals in Vital et al. 2017, are known butyrate producers (with a majority of the analyzed representative genomes containing known butyrate production pathways, as described in Table S2 of the original publication), and accounted for the majority of the total butyrate pathway abundances in human metagenomics data. We did not consider *E. ventriosum, E. hallii*, and *E. rectale* from our analyses as these species-level taxa were not present in our data and do not comprise one genus with conserved butyrate production (Table S2 from Vital et al. 2017). The genera found in each dataset and their abundances in each of the donor stool samples are plotted in Supplementary Figures 1-3. Statistical significance was assessed using the ttest_ind function from the scipy.stats Python package.

### Stool metabolomics

Metabolomics data was generated as described in Poyet, Groussin, Gibbons et al. 2019. and downloaded after personal communication with the authors. The processed data has since been made available at the NIH Common Fund’s Metabolomics Data Repository and Coordinating Center website, the Metabolomics Workbench, where it has been assigned Project ID PR000804. The data can be accessed directly via its Project DOI: 10.21228/M8RM32.

For donors with multiple samples, we considered the mean metabolite abundances across all sampled time points. We identified three short chain fatty acids (SCFAs) in the data (propionate, butyrate, and isovalerate) and the major primary (cholate and chenodeoxycholate) and secondary (deoxycholate and lithocholate) bile acids. Lithocholate abundances were available for both C-18 negative and HILIC negative modes; we considered only the C-18 negative data to match the other bile acids. Bile acid conversion rates were calculated as in Kakiyama et al. 2013. Donors were ranked based on their average SCFA abundances and based on the total bile acid conversion ratio ( (lithocholate + deoxycholate) / (chenodeoxycholate + cholate)).

### Power simulation

We performed a simulation to determine the power of FMT trials to retrospectively find associations between donor bacterial abundances and FMT response. We used case-control gut microbiome datasets from MicrobiomeHD (Duvallet et al. 2017) to model different effect sizes for FMT response. Here, we use “effect size” to mean the number of genera which are differentially abundant between patients who respond to FMT vs. patients who do not. Per the results in MicrobiomeHD, we used infectious diarrhea to model a large effect (Schubert et al. 2014), colorectal cancer to model a medium effect (Baxter et al. 2016), and obesity to model a small effect (Goodrich et al. 2014). We collapsed OTUs to genus-level as in Duvallet et al. 2017 and ranked genera according to their signal-to-noise ratio in each entire dataset, where the signal-to-noise was calculated as the difference in mean log abundance in cases and controls divided by the standard deviation of the log abundances across all samples. We considered the 10 genera with the largest absolute signal-to-noise ratios as our “top hits” in the main text.

We modeled different FMT clinical trial designs and outcomes by varying the number of total patients in the trial and the percent of FMT responders (i.e. the number of patients we selected from the original “case” group relative to the original “control” patients, to model FMT responders and non-responders). For each of these designs, we subsampled the correct number of case samples to represent FMT responders and control samples to represent non-responders from the original datasets. We identified significantly differentially abundant genera with the ‘kruskalwallis’ function from scipy.stats.mstats (scipy v. 1.1.0) as genera with q < 0.05 after multiple hypothesis testing correction with the multipletests function (method=’fdr_bh’) from the statsmodels.sandbox.stats.multicomp package (statsmodels v. 0.9.0). We then counted how many of the top genera identified through the signal-to-noise ranking were identified as significantly different as a proxy for the power to detect effects.

### Code and data availability

Code to reproduce all of these analyses and figures can be found at https://github.com/cduvallet/donor-selection/. Data were downloaded from original sources as described above.

## Data Availability

Code to reproduce all of these analyses and figures can be found at https://github.com/cduvallet/donor-selection/. Data were downloaded from original sources as described in the Methods section.

## Acknowledgments

We thank Carolyn Edelstein for her contributions on addressing safety considerations related to FMT, and Scott Olesen for his perspective on this work.

## Supplementary Figures

**Supplementary Figure 1.**
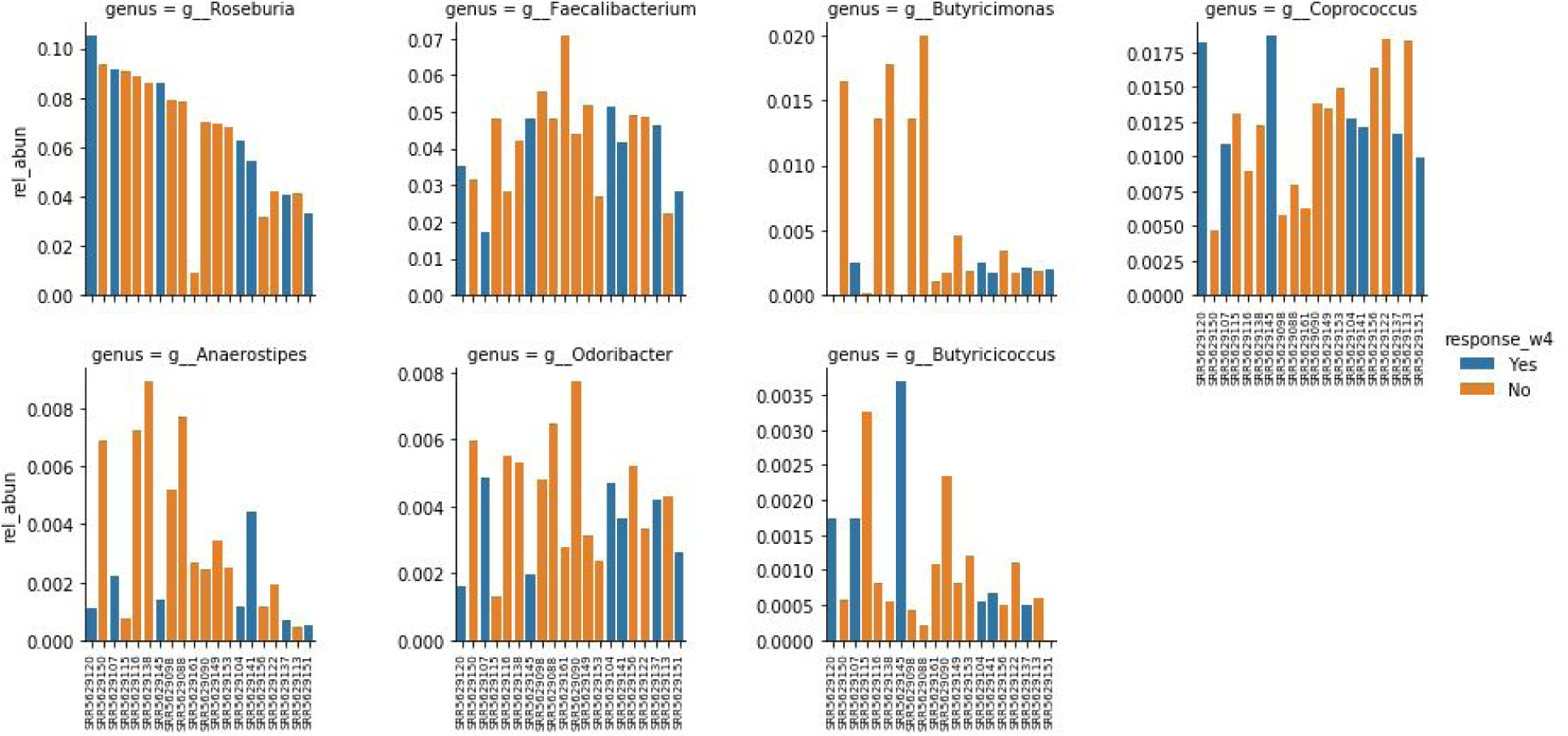
Relative abundance of butyrate-producing genera in Jacob 2017 study. Donors are ordered along the x-axis in the same order for all plots. The respective patient response status is indicated through the bar color (blue = response, orange = no response).

**Supplementary Figure 2.**
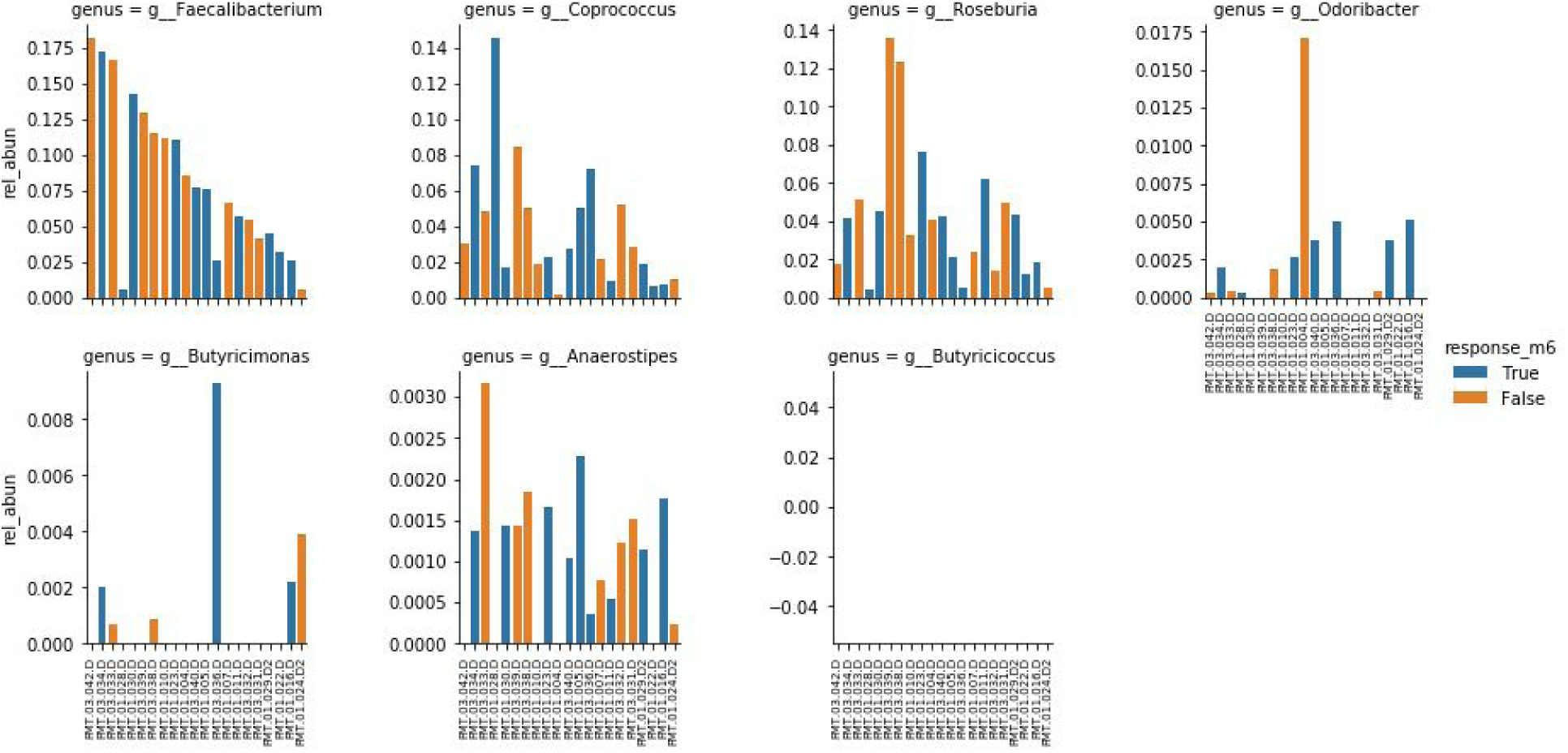
Relative abundance of butyrate-producing genera in Goyal 2018 study. Donors are ordered along the x-axis in the same order for all plots. The respective patient response status is indicated through the bar color (blue = response, orange = no response). Note: Butyricicoccus was not present in any of the donor samples.

**Supplementary Figure 3.**
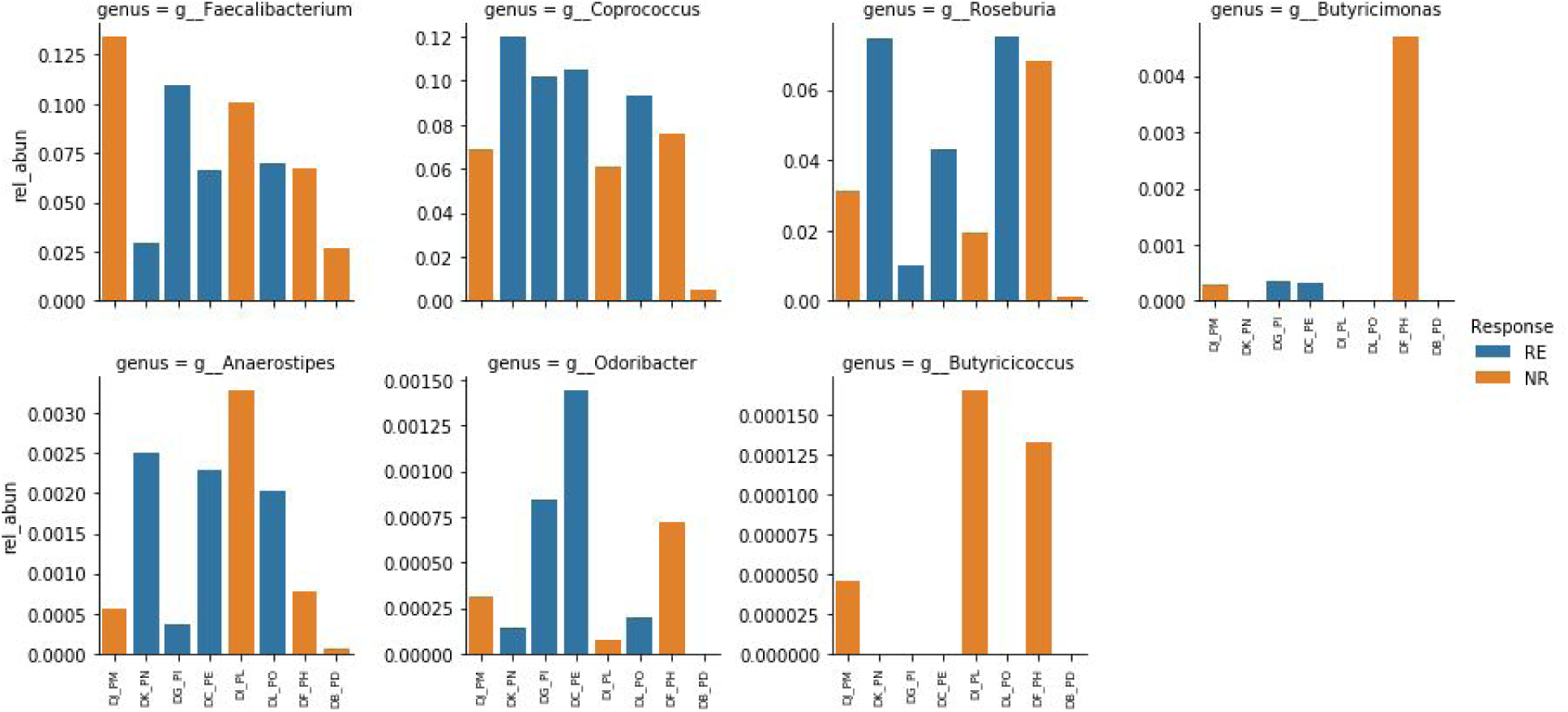
Relative abundance of butyrate-producing genera in Kump 2018 study. Donors are ordered along the x-axis in the same order for all plots. The respective patient response status is indicated through the bar color (blue = response, orange = no response).

**Supplementary Figure 4.**
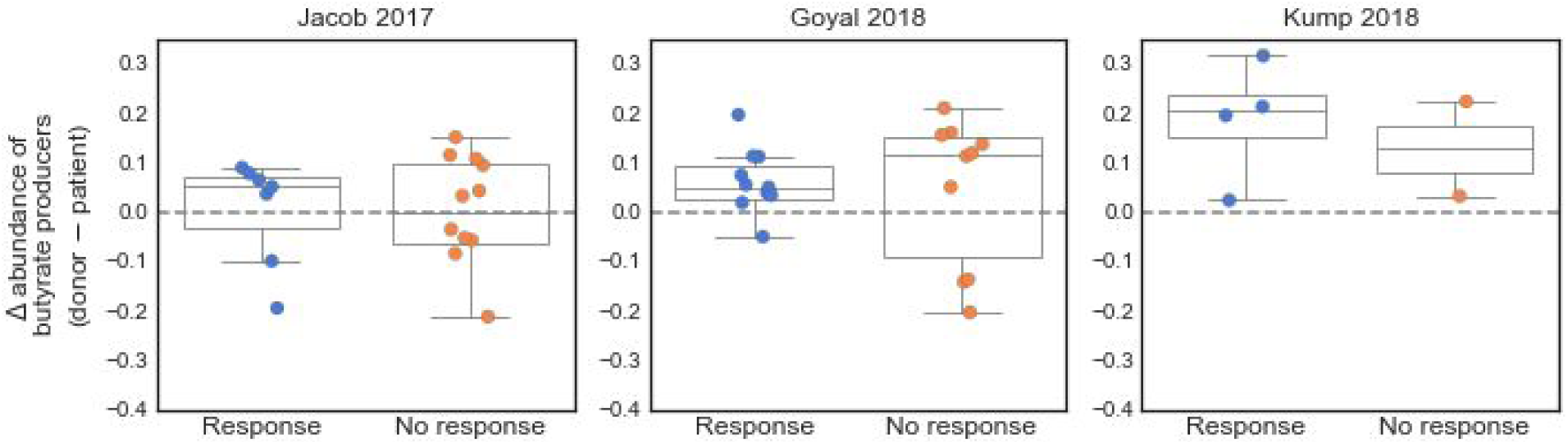
Difference between abundance of butyrate producers in donor sample and respective patient sample, stratified by patient response. The difference was calculated by subtracting the patient’s total abundance of butyrate producers from the total abundance in their respective donor sample. Butyrate producers were identified as described in the Methods.

## References

Amir, A. et al. 2017. Deblur rapidly resolves single-nucleotide community sequence patterns. mSystems, 2(2), pp. e00191–16.

Bafeta, A. et al. 2017. Methods and Reporting Studies Assessing Fecal Microbiota Transplantation: A Systematic Review. Annals of Internal Medicine, 167(1), pp. 34–39.

Bajaj, J.S., et al., 2017. Fecal microbiota transplant from a rational stool donor improves hepatic encephalopathy: a randomized clinical trial. Hepatology, 66(6), pp. 1727–1738.

Baxter, N.T., et al., 2016. Microbiota-based model improves the sensitivity of fecal immunochemical test for detecting colonic lesions. Genome medicine, 8(1), p. 37.

Bello, M.G.D., et al., 2018. Preserving microbial diversity. Science, 362(6410), pp. 33–34.

Blanton, L.V., et al., 2016. Gut bacteria that prevent growth impairments transmitted by microbiota from malnourished children. Science, 351(6275), p. aad3311.

Bokulich, N.A., et al., 2018. Optimizing taxonomic classification of marker-gene amplicon sequences with QIIME 2’s q2-feature-classifier plugin. Microbiome, 6(1), p. 90.

Bolyen E, Rideout JR, Dillon MR, Bokulich NA, et al., 2019. QIIME 2: Reproducible, interactive, scalable, and extensible microbiome data science. Nature Biotechnology, p. 1.

Britton, R. and Young, V.B., 2014. Role of the Intestinal Microbiota in Resistance to Colonization by Clostridium difficile. Gastroenterology, 146(6), pp. 1547–1553.

Bullman, S., et al., 2017. Analysis of Fusobacterium persistence and antibiotic response in colorectal cancer. Science, 358(6369), pp. 1443–1448.

Cammarota, G. et al. 2017. European consensus conference on faecal microbiota transplantation in clinical practice. Gut, 66(4), pp. 569–580.

Chen, T. et al. 2017. Fiber-utilizing capacity varies in Prevotella-versus Bacteroides-dominated gut microbiota. Scientific reports, 7(1), p. 2594.

Chu, N.D., et al., 2019. Dynamic colonization of microbes and their functions after fecal microbiota transplantation for inflammatory bowel disease. bioRxiv, p. 649384.

Costello, S.P. et al. 2017. Short Duration, Low Intensity, Pooled Fecal Microbiota Transplantation Induces Remission in Patients with Mild-Moderately Active Ulcerative Colitis: A Randomised Controlled Trial. Gastroenterology, 152(5), pp. S198–S199.

Cox, L.M. et al. 2014. Altering the intestinal microbiota during a critical developmental window has lasting metabolic consequences. Cell, 158(4), pp. 705–721.

De Filippo, C. et al. 2010. Impact of diet in shaping gut microbiota revealed by a comparative study in children from Europe and rural Africa. Proceedings of the National Academy of Sciences of the United States of America, 107(33), pp. 14691–14696.

Duvallet, C., et al., 2017. Meta-analysis of gut microbiome studies identifies disease-specific and shared responses. Nature communications, 8(1), p. 1784.

Friedman, J., & Alm, E. J. 2012. Inferring correlation networks from genomic survey data. PLoS computational biology, 8(9), e1002687.

Food and Drug Administration, 2013. Enforcement Policy Regarding Investigational New Drug Requirements for Use of Fecal Microbiota for Transplantation to Treat Clostridium difficile Infection Not Responsive to Standard Therapies. Accessed July 22, 2019. https://www.fda.gov/media/86440/download

Food and Drug Administration, 2019. Information Pertaining to Additional Safety Protections Regarding Use of Fecal Microbiota for Transplantation – Screening and Testing of Stool Donors for Multi-drug Resistant Organisms. Accessed July 22, 2019. https://www.fda.gov/vaccines-blood-biologics/safety-availability-biologics/information-pertaining-additional-safety-protections-regarding-use-fecal-microbiota-transplantation

Gelfand, 2018. Fecal Microbiota Transplantation (FMT) of FMP30 in Relapsing-Remitting Multiple Sclerosis - Full Text View - ClinicalTrials.gov. ClinicalTrials.gov. Available at: https://clinicaltrials.gov/ct2/show/NCT03594487 [Accessed November 28, 2018].

Goodrich, J.K., et al., 2014. Human genetics shape the gut microbiome. Cell, 159(4), pp. 789–799.

Goyal, A., et al., 2018. Safety, clinical response, and microbiome findings following fecal microbiota transplant in children with inflammatory bowel disease. Inflammatory bowel diseases, 24(2), pp. 410–421.

Halfvarson, J., et al., 2017. Dynamics of the human gut microbiome in inflammatory bowel disease. Nature microbiology, 2(5), p. 17004.

Hofmann, A.F., 1989. Enterohepatic circulation of bile acids. Handbook of Physiology. The Gastrointestinal System, 4, pp. 567–596.

Hsiao, A., et al., 2014. Members of the human gut microbiota involved in recovery from Vibrio cholerae infection. Nature, 515(7527), p. 423.

The Human Microbiome Project Consortium. (2012). Structure, function and diversity of the healthy human microbiome. nature, 486(7402), 207.

Jacob, V., et al., 2017. Single delivery of high-diversity fecal microbiota preparation by colonoscopy is safe and effective in increasing microbial diversity in active ulcerative colitis. Inflammatory bowel diseases, 23(6), pp. 903–911.

Kakiyama, G., et al., 2013. Modulation of the fecal bile acid profile by gut microbiota in cirrhosis. Journal of hepatology, 58(5), pp. 949–955.

Kelly, C.R. et al. 2016. Effect of Fecal Microbiota Transplantation on Recurrence in Multiply Recurrent Clostridium difficile Infection: A Randomized Trial. Annals of internal medicine, 165(9), pp. 609–616.

Koeth, R.A., et al., 2013. Intestinal microbiota metabolism of L-carnitine, a nutrient in red meat, promotes atherosclerosis. Nature medicine, 19(5), p. 576.

Kootte, R.S. et al. 2017. Improvement of Insulin Sensitivity after Lean Donor Feces in Metabolic Syndrome Is Driven by Baseline Intestinal Microbiota Composition. Cell metabolism, 26(4), pp. 611–619.e6.

Kostic, A.D., et al., 2013. Fusobacterium nucleatum potentiates intestinal tumorigenesis and modulates the tumor-immune microenvironment. Cell host & microbe, 14(2), pp. 207–215.

Kostic, A.D. et al. 2015. The dynamics of the human infant gut microbiome in development and in progression toward type 1 diabetes. Cell host & microbe, 17(2), pp. 260–273.

Kump, P., et al., 2018. The taxonomic composition of the donor intestinal microbiota is a major factor influencing the efficacy of faecal microbiota transplantation in therapy refractory ulcerative colitis. Alimentary pharmacology & therapeutics, 47(1), pp. 67–77.

Louis, P., et al., 2010. Diversity of human colonic butyrate-producing bacteria revealed by analysis of the butyryl-CoA: acetate CoA-transferase gene. Environmental microbiology, 12(2), pp. 304–314.

McDonald, L.C. et al. 2018. Clinical Practice Guidelines for Clostridium difficile Infection in Adults and Children: 2017 Update by the Infectious Diseases Society of America (IDSA) and Society for Healthcare Epidemiology of America (SHEA). Clinical infectious diseases: an official publication of the Infectious Diseases Society of America, 66(7), pp. e1–e48.

Moayyedi, P. et al. 2015. Fecal Microbiota Transplantation Induces Remission in Patients With Active Ulcerative Colitis in a Randomized Controlled Trial. Gastroenterology, 149(1), pp. 102–109.e6.

van Nood, E., Dijkgraaf, M.G.W. & Keller, J.J., 2013. Duodenal infusion of feces for recurrent Clostridium difficile. The New England journal of medicine, 368(22), p. 2145.

Olesen, S., Gurry, T., and Alm, E.J., 2017. Designing fecal microbiota transplant trials that account for differences in donor stool efficacy. Statistical methods in medical research, 27(10), pp. 2906–2917.

Olesen, S.W., et al., 2018. Searching for superstool: maximizing the therapeutic potential of FMT. Nature reviews. Gastroenterology & hepatology, 15(7), pp. 387–388

OpenBiome, 2019. “OpenBiome Quality & Safety Program.” https://www.openbiome.org/safety. Accessed August 9, 2019.

Osman, M., et al., 2016, December. Donor efficacy in fecal microbiota transplantation for recurrent Clostridium difficile: evidence from a 1,999-Patient Cohort. In Open Forum Infectious Diseases (Vol. 3, No. suppl_1). Oxford University Press.

Osman, M., 2018. Transfer of Healthy Gut Flora for Restoration of Intestinal Microbiota Via Enema for Severe Acute Malnutrition - Full Text View - ClinicalTrials.gov. Available at: https://clinicaltrials.gov/ct2/show/NCT03087097 [Accessed November 28, 2018].

Ott, S.J. et al. 2017. Efficacy of Sterile Fecal Filtrate Transfer for Treating Patients With Clostridium difficile Infection. Gastroenterology, 152(4), pp. 799–811.e7.

Ou, J. et al. 2013. Diet, microbiota, and microbial metabolites in colon cancer risk in rural Africans and African Americans. The American journal of clinical nutrition, 98(1), pp. 111–120.

Panchal, P. et al. 2018. Scaling Safe Access to Fecal Microbiota Transplantation: Past, Present, and Future. Current gastroenterology reports, 20(4), p. 14.

Paramsothy, S. et al. 2017. Multidonor intensive faecal microbiota transplantation for active ulcerative colitis: a randomised placebo-controlled trial. The Lancet, 389(10075), pp. 1218–1228.

Poyet, M., Groussin, M., Gibbons, SM., et al. 2019. A library of human gut bacterial isolates paired with longitudinal multiomics data enables mechanistic microbiome research. Nature Medicine (in press).

Quraishi, M.N. et al. 2017. Systematic review with meta-analysis: the efficacy of faecal microbiota transplantation for the treatment of recurrent and refractory Clostridium difficile infection. Alimentary pharmacology & therapeutics, 46(5), pp. 479–493.

Rabesandratana, T., 2018. ‘Poop vault’ of human feces could preserve gut’s microbial biodiversity--and help treat disease. Science, Nov. 2018.

Rubinstein, M.R., et al., 2013. Fusobacterium nucleatum promotes colorectal carcinogenesis by modulating E-cadherin/β-catenin signaling via its FadA adhesin. Cell host & microbe, 14(2), pp. 195–206.

Scheppach, W., et al., 1992. Effect of butyrate enemas on the colonic mucosa in distal ulcerative colitis. Gastroenterology, 103(1), pp. 51–56.

Schirmer, M., et al., 2018. Compositional and Temporal Changes in the Gut Microbiome of Pediatric Ulcerative Colitis Patients Are Linked to Disease Course. Cell host & microbe, 24(4), pp. 600–610.

Schubert, A.M., et al., 2014. Microbiome data distinguish patients with Clostridium difficile infection and non-C. difficile-associated diarrhea from healthy controls. MBio, 5(3), pp. e01021–14.

Shade, A. and Handelsman, J., 2012. Beyond the Venn diagram: the hunt for a core microbiome. Environmental microbiology, 14(1), pp. 4–12.

Stein, R.R., et al., 2018. Computer-guided design of optimal microbial consortia for immune system modulation. eLife, 7, p. e30916.

Stokholm, J. et al. 2018. Maturation of the gut microbiome and risk of asthma in childhood. Nature communications, 9(1), p. 141.

Subramanian, S., et al., 2014. Persistent gut microbiota immaturity in malnourished Bangladeshi children. Nature, 510(7505), p. 417.

Surawicz, C.M. et al. 2013. Guidelines for diagnosis, treatment, and prevention of Clostridium difficile infections. The American journal of gastroenterology, 108(4), pp. 478–98; quiz 499.

Vital, M., et al., 2017. Colonic Butyrate-Producing Communities in Humans: an Overview Using Omics Data. MSystems, 2(6), pp. e00130–17.

Wang, X. and Gibson, G.R. 1993. Effects of the in vitro fermentation of oligofructose and inulin by bacteria growing in the human large intestine. Journal of applied microbiology, 75(4), pp. 373–380.

Wang, Z., et al., 2015. Non-lethal inhibition of gut microbial trimethylamine production for the treatment of atherosclerosis. Cell, 163(7), pp. 1585–1595.

Wilck, N., et al., 2017. Salt-responsive gut commensal modulates T H 17 axis and disease. Nature, 551(7682), p. 585.

Wilson, B. C., Vatanen, T., Cutfield, W. S., & O’Sullivan, J. M. (2019). The Super-Donor Phenomenon in Fecal Microbiota Transplantation. Front. Cell. Infect. Microbiol. 9: 2. doi: 10.3389/fcimb.

Yatsunenko, T. et al. 2012. Human gut microbiome viewed across age and geography. Nature, 486(7402), pp. 222–227.

Zaneveld, J., et al., 2017. Stress and stability: applying the Anna Karenina principle to animal microbiomes. Nature microbiology, 2(9), p. 17121.

Zuo, T. et al. 2018. Bacteriophage transfer during faecal microbiota transplantation in Clostridium difficile infection is associated with treatment outcome. Gut, 67(4), pp. 634–643.

